# Safety Monitoring of Mpox Vaccines in Adults Aged 18–64 Years during the 2022 Mpox Outbreak in the U.S.

**DOI:** 10.1101/2024.12.17.24319168

**Authors:** Joann F. Gruber, Kathryn Matuska, Carla E. Zelaya, Jing Wang, Cheryl N McMahill-Walraven, Lauren Peetluk, Alex Secora, Shwetha Krishnakumar, Mao Hu, Kandace L. Amend, Jennifer Song, Daniel C. Beachler, Patricia C. Lloyd, Yoganand Chillarige, Jane A. Gwira, Wafa Tarazi, Tainya C. Clarke, Djeneba Audrey Djibo, Richard A. Forshee, Azadeh Shoaibi, Steven A. Anderson

## Abstract

Two vaccines (JYNNEOS; ACAM2000) were available in the United States to prevent mpox during the 2022 clade II mpox outbreak with the majority of people receiving JYNNEOS. As part of the Biologics Effectiveness and Safety (BEST) Initiative, FDA monitored the safety of JYNNEOS using three commercial health plan claims databases supplemented with local and state Immunization Information System data. We assessed vaccine uptake, described the vaccinated population, monitored rates of 11 potential adverse events (AEs) following vaccination, and compared the observed AE rates to expected background rates. There were 152,001 JYNNEOS doses administered. Of the 92,340 people receiving at least one JYNNEOS dose, most were males (93.1%), aged 25–44 years (63.3%), lived in urban areas (98.7%), and 64.2% received a second dose. AE rates following JYNNEOS doses were similar to expected background rates. Additional safety studies of larger vaccinated populations may be needed to evaluate rare AEs including myocarditis/pericarditis.

## 1. Introduction

In 2022, a global outbreak of clade II mpox spread to areas without historically reported cases [1]. In the United States (U.S.), the first case was reported in May 2022. As of January 2024, there have been over 32,000 reported cases and 58 deaths in the U.S. [2]. While most of the cases occurred during the summer and fall of 2022, clade II mpox cases continue to be reported at low levels in the U.S. and other countries [3, 4].

Two orthopoxvirus vaccines (JYNNEOS and ACAM2000) were made available for the prevention of mpox in the U.S. during the 2022 outbreak. At the start of the outbreak, the Centers for Disease Control and Prevention (CDC) recommended vaccination for those with known or presumed exposure to mpox [5]; by the end of September 2022, CDC expanded recommendations to pre-exposure vaccination for those at highest risk [6]. The JYNNEOS vaccine is a two-dose vaccine (4 weeks apart) containing the Modified Vaccinia Ankara-Bavarian Nordic (MVA-BN) strain—a live, attenuated, non-replicating orthopoxvirus [7]—and considered safer for use in immunocompromised persons than other live vaccines [6]. It was licensed by the U.S. Food and Drug Administration (FDA) in 2019 for the prevention of mpox and smallpox in adults aged 18 years and older. The ACAM2000 vaccine contains a live, attenuated, replicating strain of the vaccinia virus and was initially licensed by FDA in 2007 for the prevention of smallpox only [8]. It was made available for use to prevent mpox under the Expanded Access Investigational New Drug mechanism [9]. However, ACAM2000 was not recommended for use in all populations during the 2022 clade II outbreak and was available only by special request [5].

There have been limited active and passive safety surveillance studies of the JYNNEOS vaccine published globally [10–12]. Large cohort-based safety studies assessing several potential adverse events (AEs) of interest have not been published to date.

As part of the Biologics Effectiveness and Safety (BEST) Initiative, FDA assessed the safety of JYNNEOS vaccines used during the 2022 clade II mpox outbreak by estimating the incidence rates of 11 potential adverse events (AEs) following JYNNEOS vaccination among U.S. adults aged 18–64 years using large U.S. based commercial health insurance claims databases supplemented with vaccination data from Immunization Information Systems (IIS) and compared these rates to the age-sex standardized expected rates for the vaccinated population.

## 2. Methods

Data from three U.S. commercial claims databases in the FDA BEST Initiative were used: Carelon Research (Elevance Health, including Anthem, and affiliated health plans), CVS Health (Aetna and affiliated health plans), and Optum (commercial health plans). To increase the capture of vaccines for people enrolled in these commercial insurance plans, claims data were supplemented with available local and state IIS data. The number of state IIS jurisdictions varied by database (CVS Health: 23, Carelon Research: 10, Optum: 6; Supplementary Table 1). Data were refreshed monthly to conduct near-real time surveillance as vaccines were administered. Our study population included individuals who received at least one dose of JYNNEOS and were aged 18–64 years at the time of vaccination. The study period started on May 10, 2022 and ended on July 4, 2023 (Carelon Research), June 30, 2023 (CVS Health), and July 8, 2023 (Optum). Exposure was defined as receipt of any dose of JYNNEOS, as identified by product codes such as Current Procedural Terminology/Healthcare Common Procedure Coding System codes, National Drug Codes, and CVX codes in any care setting in claims or IIS data [13]. Since there was extremely limited use of ACAM2000 (N = 239 doses across the three databases), our monitoring focused on JYNNEOS only.

We assessed the count and trend of JYNNEOS doses over time. In addition, we summarized the distribution of demographic and geographic characteristics of the JYNNEOS vaccinated population and the percentage of people receiving a complete JYNNEOS vaccine series (i.e., at least two doses).

We monitored the occurrence of 11 potential AEs in outcome-specific risk windows following JYNNEOS vaccination. Potential AEs included serious AEs following other vaccinations including those known or possibly related to smallpox vaccination. To be included in an AE-specific analysis, persons were required to be continuously enrolled in the health plan from the start of the AE-specific clean window through the date of vaccination, and must not have had the specific AE during the clean window. The list of AEs, pre-vaccination clean windows, and post-vaccination risk windows are detailed in Table 1. Claims-based AE algorithms were developed based on literature review and in consultation with clinical experts [13]. Observed rates per 100,000 person years of each AE following any JYNNEOS dose and corresponding exact 95% confidence intervals (CIs) were calculated [14]. Follow-up time continued from the start of the AE-specific risk window until a person experienced an AE, death, disenrollment, a third JYNNEOS or second ACAM2000 dose, or the end of the risk window. We aggregated all statistics across the three databases to protect privacy. To contextualize the observed rates, we estimated the age-sex standardized expected rates of each AE and 95% CIs [15] by applying approximate age-sex specific 2019 background rates (where available) (Supplementary Table 2), estimated using methods from prior work [16], to the age-sex distribution of the JYNNEOS-vaccinated population.

**Table 1.** List of clean windows and risk windows used for safety monitoring of JYNNEOS vaccination, by adverse event and healthcare setting, in U.S. adults aged 18–64 years.

| **Adverse Event** | **Healthcare Setting<sup>a</sup>** | **Clean Window (days)<sup>b</sup>** | **Risk Window (days)** |
| --- | --- | --- | --- |
| Acute Myocardial Infarction | IP | 365 | 1–28 |
| Anaphylaxis | IP, OP-ED | 30 | 0–1 |
| Bell's Palsy | IP, OP/PB | 183 | 1–42 |
| Cardiomyopathy | IP, OP-ED | 365 | 1–42 |
| Deep Vein Thrombosis | IP, OP/PB | 365 | 1–28 |
| Encephalitis, Myelitis, or Encephalomyelitis | IP | 183 | 1–42 |
| Guillain-Barré Syndrome | IP-primary position only | 365 | 1–42 |
| Myocarditis/Pericarditis | IP, OP-ED | 365 | 1–21 & 1–42 |
| Non-hemorrhagic Stroke | IP | 365 | 1–28 |
| Pulmonary Embolism | IP | 365 | 1–28 |
| Transverse Myelitis | IP, OP-ED | 365 | 1–42 |

<sup>a</sup> Care setting definitions: IP refers to inpatient facility claims. IP-primary position only refers to only those inpatient claims which are the primary record on the claim. OP-ED refers to a subset of outpatient facility claims occurring in the emergency department. OP/PB refers to all outpatient facility claims, and professional/provider claims except those with a laboratory place of service.
<sup>b</sup> In order to be considered an incident occurrence of an AE, it must not be observed in a pre-specified period prior to vaccination.

To investigate potential pre-existing comorbidities that could influence the risk of myocarditis/pericarditis following vaccination, we completed a review of all claims in the 365 days prior and 105 days following the myocarditis/pericarditis diagnosis that occurred 1–42 days following JYNNEOS vaccination. For those with available data, a clinician reviewed all claims to determine if persons had any pre-existing conditions for myocarditis/pericarditis.

This surveillance activity was conducted as part of the FDA public health surveillance mandate [17].

## 3. Results

A total of 152,001 JYNNEOS doses were observed with the majority administered between July and October of 2022 (Figure 1). Of the 92,340 people vaccinated with at least one JYNNEOS dose, most were male (93.1%), 25–44 years of age at the time of first dose (63.3%), lived in urban areas (98.7%) and resided in Health and Human Services regions two (32.4%) and nine (32.5%) (Table 2). Among those who received a first dose of JYNNEOS, 64.2% received a second dose, but this differed by sex and age group (Supplementary Table 3).

**Figure 1.**
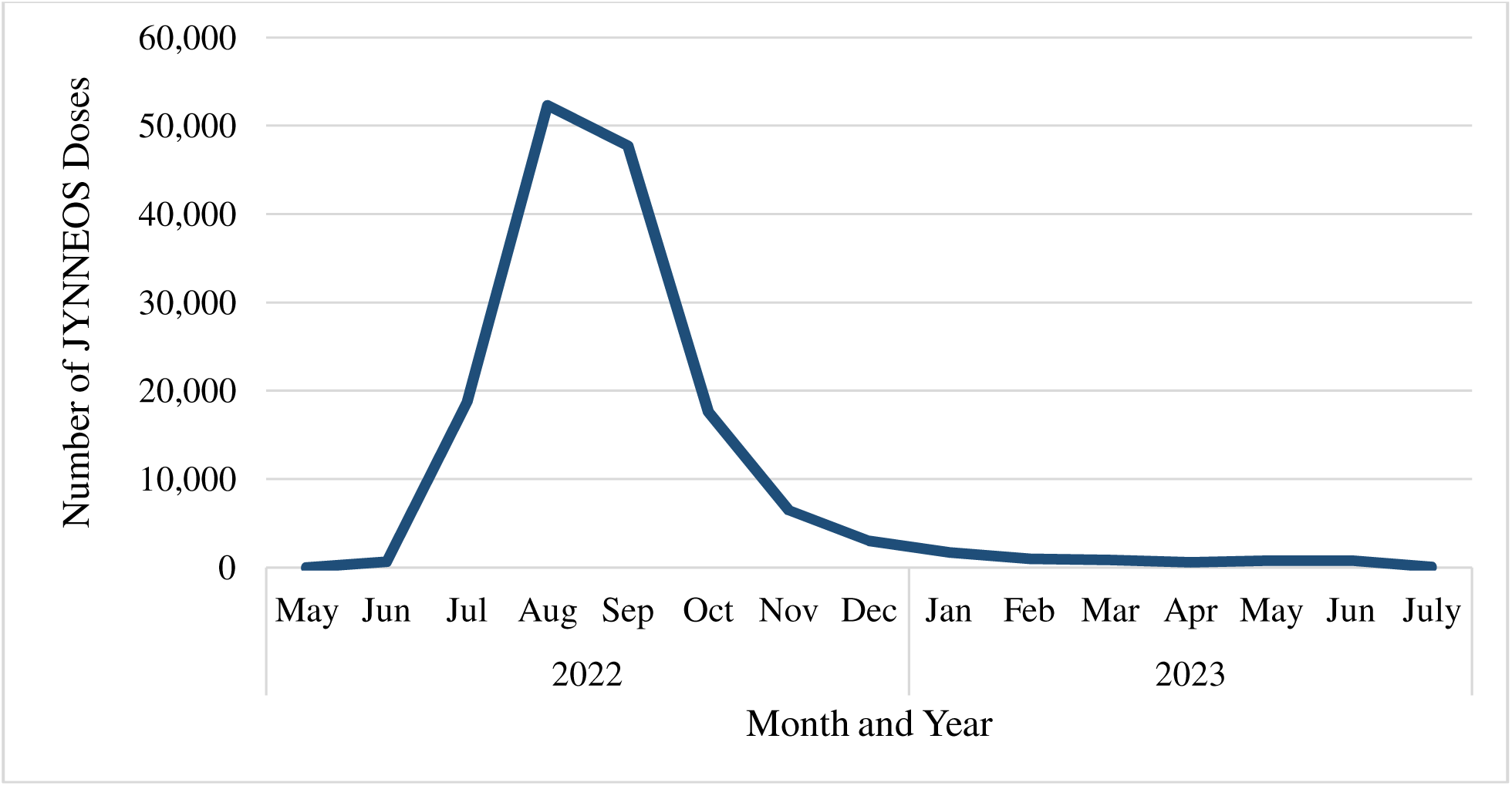
Monthly counts of JYNNEOS doses administered to U.S. adults aged 18–64 years from May 2022 through July 2023.

**Table 2.** Demographic and geographic characteristics of U.S. adults aged 18–64 years who received at least one JYNNEOS dose.

| Characteristic | N | % |
| --- | --- | --- |
| **Total** | 92,340 | 100 |
| **Sex** |  |  |
| Female | 6,243 | 6.8 |
| Male | 85,947 | 93.1 |
| Missing/Unknown | 150 | 0.2 |
| **Age (years)** |  |  |
| 18–24 | 6,635 | 7.2 |
| 25–34 | 33,269 | 36.0 |
| 35–44 | 25,202 | 27.3 |
| 45–54 | 15,344 | 16.6 |
| 55–64 | 11,890 | 12.9 |
| **HHS Region** |  |  |
| Region 1: CT, ME, NH, RI, VT | 2,810 | 3.0 |
| Region 2: NJ, NY, PR, VI | 29,923 | 32.4 |
| Region 3: DE, DC, MD, PA, VA, WV | 3,532 | 3.8 |
| Region 4: AL, FL, GA, KY, MS, NC, SC, TN | 9,780 | 10.6 |
| Region 5: IL, IN, MI, MN, OH, WI | 6,474 | 7.0 |
| Region 6: AR, LA, NM, OK, TX | 4,708 | 5.1 |
| Region 7: IA, KS, MO, NE | 356 | 0.4 |
| Region 8: CO, MT, ND, SD, UT, WY | 2,075 | 2.2 |
| Region 9: AZ, CA, HI, NV, AS, MP, FM, GU, MH, PW | 30,045 | 32.5 |
| Region 10: AK, ID, OR, WA | 2,598 | 2.8 |
| Missing/Unknown | 39 | 0.0 |
| **Urban/Rural** |  |  |
| Urban | 91,097 | 98.7 |
| Rural | 1,180 | 1.3 |
| Missing/Unknown | 63 | 0.1 |

There were 15 or fewer cases of each AE, and AE rates ranged from zero to 188.2 events per 100,000 person-years (Figure 2). Historical background rates from 2019 were available for 9 of the 11 AEs; background rates were not available for cardiomyopathy or the composite outcome of encephalitis, myelitis, or encephalomyelitis. There was overlap between the 95% CIs of observed and age-sex standardized expected rates. Point estimates of age-sex standardized expected background rates were higher than observed rates in all instances except two (myocarditis/pericarditis and anaphylaxis). However, observed rates of both outcomes were imprecise and based on 10 or fewer cases. For myocarditis/pericarditis, the rate in the 1–42 days following vaccination (N = 10 cases) was 90.2 (95% CI: 43.3, 165.9) per 100,000 person-years compared to the age-sex standardized expected rate of 39.7 (95% CI: 25.7, 50.6) per 100,000 person-years. Of the ten persons with myocarditis/pericarditis in the 1–42 days following a JYNNEOS dose, claims profile information was available for eight. Of the eight people, one had myocarditis and seven had pericarditis. Based on the claims profile review, the clinician determined that all eight people had at least one risk factor for myocarditis/pericarditis present prior to vaccination, including recent coronary artery bypass grafting, alcohol abuse, atrial fibrillation, HIV, or cardiomyopathy. For anaphylaxis, the rate was elevated, but extremely imprecise, due to very small case counts.

**Figure 2.**
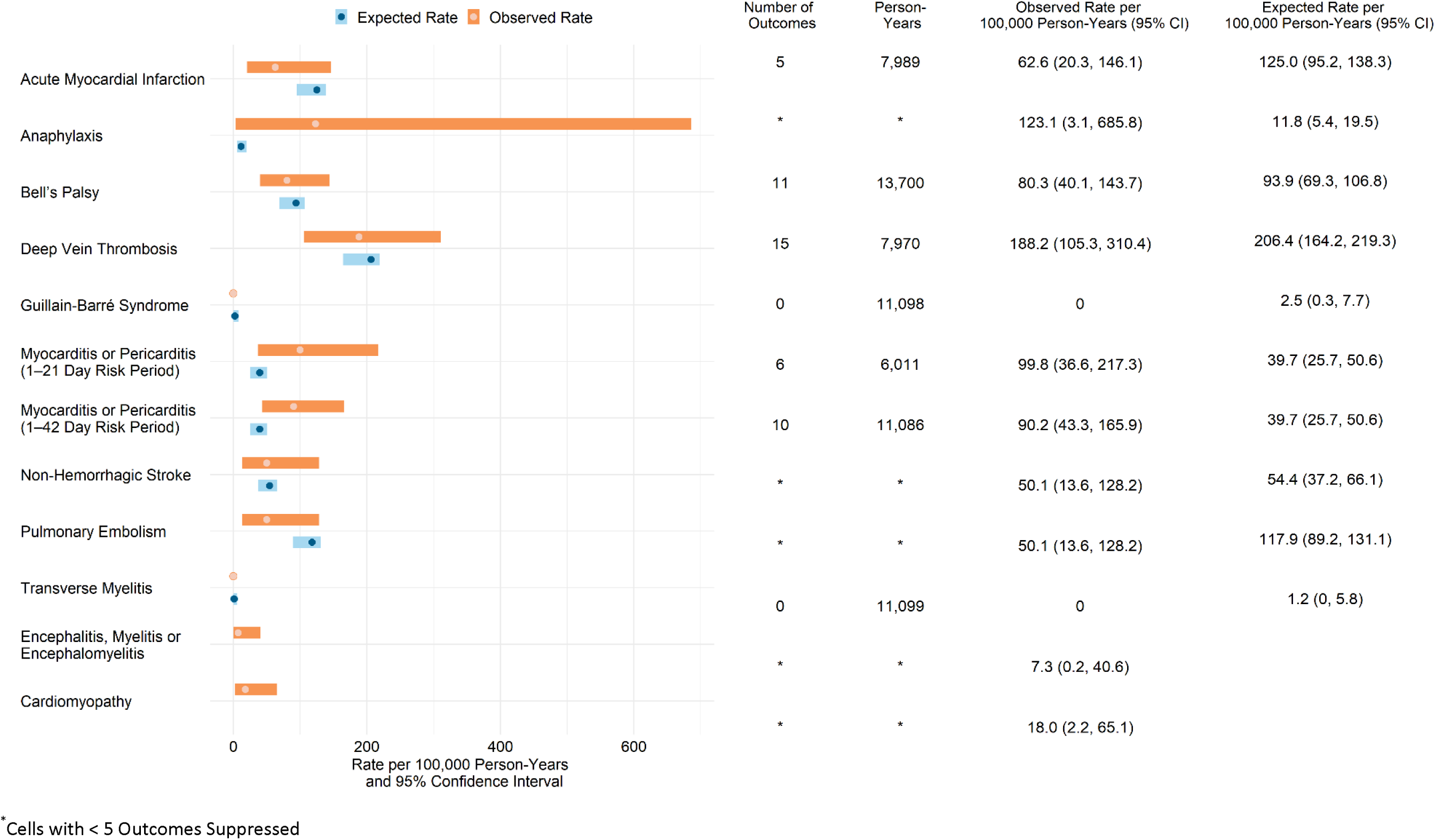
Observed and age-sex standardized expected background rates of adverse events following JYNNEOS doses administered to U.S. adults aged 18–64 years.

## 4. Discussion

This active safety surveillance study was a large post-marketing study of JYNNEOS-vaccinated persons. Using U.S. commercial claims databases supplemented with IIS data, we monitored 11 AEs following approximately 150,000 JYNNEOS doses administered to persons aged 18–64 years. We found relatively low counts and rates of AEs following JYNNEOS vaccinations. Rates were similar to age-sex standardized expected background rates for the nine outcomes with prior data available.

Myocarditis and pericarditis have been associated with receipt of replicating smallpox vaccines including Dryvax (10.4 events per 1,000 vaccinations) and ACAM2000 (5.7 events per 1,000 vaccinations) [8]. JYNNEOS is a non-replicating vaccine not known to be associated with myocarditis/pericarditis. However, JYNNEOS vaccines were not used widely prior to the 2022 clade II mpox outbreak, so safety data are limited. In our study, there were very few claims-based cases of myocarditis/pericarditis observed following JYNNEOS-vaccination (i.e., 6 cases 1–21 days and 4 cases 22–42 days following vaccination). The Vaccine Safety Datalink, a CDC active surveillance system, reported 43,253 JYNNEOS doses during the 2022 clade II mpox outbreak and reported one case of myocarditis among vaccinated males in the 30 days following each dose of the vaccine (rate per million doses (95% CI): 39 (0.1, 217.1) and 67 (1.7, 374.4) for doses 1 and 2, respectively) [12]. A retrospective review of 3,235 JYNNEOS doses administered to 2,126 individuals aged 12 years and older who received at least one JYNNEOS dose reported 10 cardiac AEs (rate of 3.1 per 1,000 doses (95% CI: 1.5, 5.7)) in the 21 days following dose receipt, but no cases of myocarditis or pericarditis [18]. Our observed rates of myocarditis/pericarditis were based off a limited number of cases and were very imprecise, making it difficult to compare them to the age-sex standardized expected background rate. The data available indicate any differences may be a result of random variability. However, since myocarditis/pericarditis rates could possibly be elevated, we reviewed additional claims data for all persons with myocarditis/pericarditis (8 of 10 available) to see if persons had other clinical factors (see “Results”) that could be associated with myocarditis/pericarditis. We found all persons had at least one pre-existing risk factor for myocarditis/pericarditis prior to vaccination. Based on these data, there is insufficient information to conclude there is increased risk of myocarditis/pericarditis following JYNNEOS vaccination.

Our study had several strengths. To our knowledge, this study was the largest post-marketing safety study of the JYNNEOS vaccine. The study population included adults enrolled in commercial insurance captured in large claims databases that collectively cover every U.S. state. During the study period, vaccines were only available through the Strategic National Stockpile, and largely administered through community-based vaccination initiatives, so they often did not generate a health insurance claim. Supplementing our claims data with data from several IIS was critical and made it possible to conduct this safety study.

Our study also had some limitations. Our age-sex standardized expected rates of AEs were based on age-sex specific background rates from the age-sex groupings from the general commercially insured population from 2019. These rates provided a general estimate for comparison; however, they may not represent the true expected background AE rates for the vaccinated population. Before September 2022, vaccines were recommended by CDC for populations at the highest risk of mpox [6]. This population may have experienced different background rates of AEs because of higher levels of comorbidities, including HIV [19]. In addition, the time period and age categories used to estimate age-sex standardized expected background rates differed slightly from the present study. These differences could also influence the accuracy of the age-sex standardized expected rates. There were no available 2019 background rates for cardiomyopathy or the composite outcome of encephalitis, myelitis, or encephalomyelitis, so we were unable to compare the observed rates to background rates. Finally, outcomes were identified using claims-based algorithms and were not chart-confirmed.

As part of the FDA BEST Initiative, this study used real-world data, including claims supplemented with IIS data, to monitor the uptake and safety of vaccines used to prevent mpox among U.S. adults aged 18–64 years during the 2022 clade II mpox outbreak. No significant safety concerns were identified following the receipt of JYNNEOS vaccines. Additional safety studies of larger vaccinated populations may be needed in the future to evaluate the potential for rare AEs including myocarditis/pericarditis.

## Data Availability

The study protocol was publicly posted as referenced in the manuscript before data analyses, and related documents can be made available where needed by contacting the corresponding author. De-identified participant data will not be shared without approval from the data partners.

## Acknowledgments

We would like to thank Cheuk Yi Lau (Acumen), Natalie Sisto (Acumen), Wenya Grace Yang (Optum Serve Consulting), Alexandra Stone (Optum Serve Consulting), Emily Myers (Optum Serve Consulting), Nwanneamaka Ume (Optum Serve Consulting), Megan Ketchell (Optum Serve Consulting), Kathryn Federici (Optum Serve Consulting), Vaibhav Sharma (CVS Health), Smita Bhatia (CVS Health), Carla Brannan (CVS Health), Ana Martinez-Baquero (CVS Health), Aparna Srikanti (CVS Health), Nancy Shaik (CVS Health), Ruth Weed (IQVIA), Rosenie Thelus (IQVIA), Nerissa Williams (IQVIA), Michael Goodman (IQVIA), Shiva Vojjala (Carelon Research), Ramya Avula (Carelon Research), Shiva Chaudhary (Carelon Research), Shanthi P Sagare (Carelon Research), Ramin Riahi (Carelon Research), Lauren Parlett (Carelon Research), Grace Stockbower (Carelon Research), Douglas Rouse (FDA), Henry Zhang (FDA), and Krista Fekecs (FDA).

## Supplementary Data for Safety Monitoring of Mpox Vaccines in Adults Aged 18–64 Years during the 2022 Mpox Outbreak in the U.S.

**Supplementary Table 1.** List of state Immunization Information System jurisdictions that were used to supplement JYNNEOS vaccination data in at least one of the commercial claims databases.

| State IIS Jurisdictions |
| --- |
| Arizona |
| California<sup>a</sup> |
| Colorado |
| Delaware |
| Georgia |
| Illinois<sup>a</sup> |
| Indiana |
| Kentucky |
| Maryland |
| Massachusetts |
| Michigan |
| Minnesota |
| Nevada |
| New Jersey |
| New Mexico |
| New York<sup>a</sup> |
| North Carolina |
| Oklahoma |
| Pennsylvania<sup>a</sup> |
| Texas |
| Vermont |
| Virginia |
| Washington |
| Wisconsin |

<sup>a</sup> May or may not include local IIS jurisdictions within that state.

**Supplementary Table 2.**
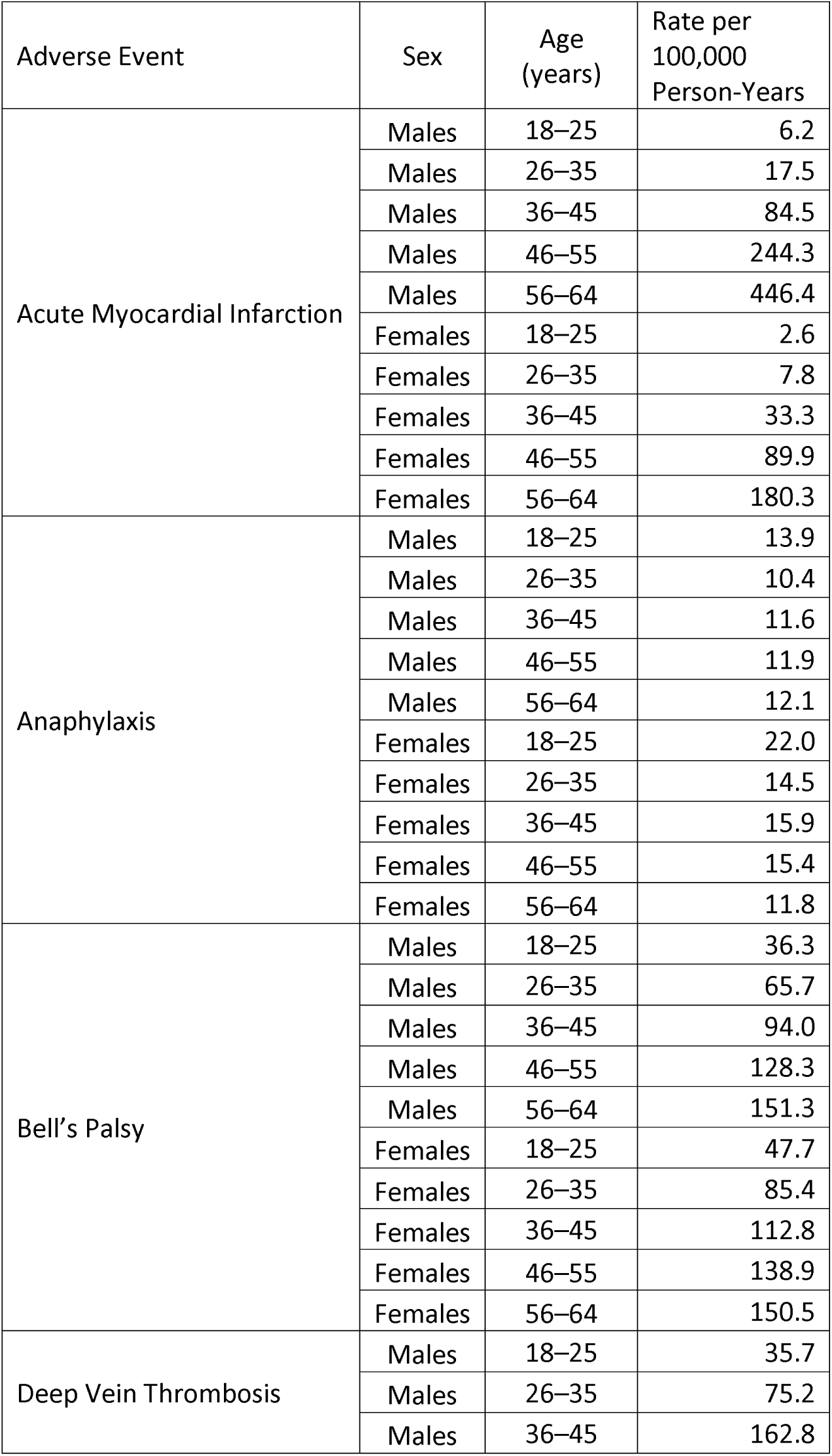
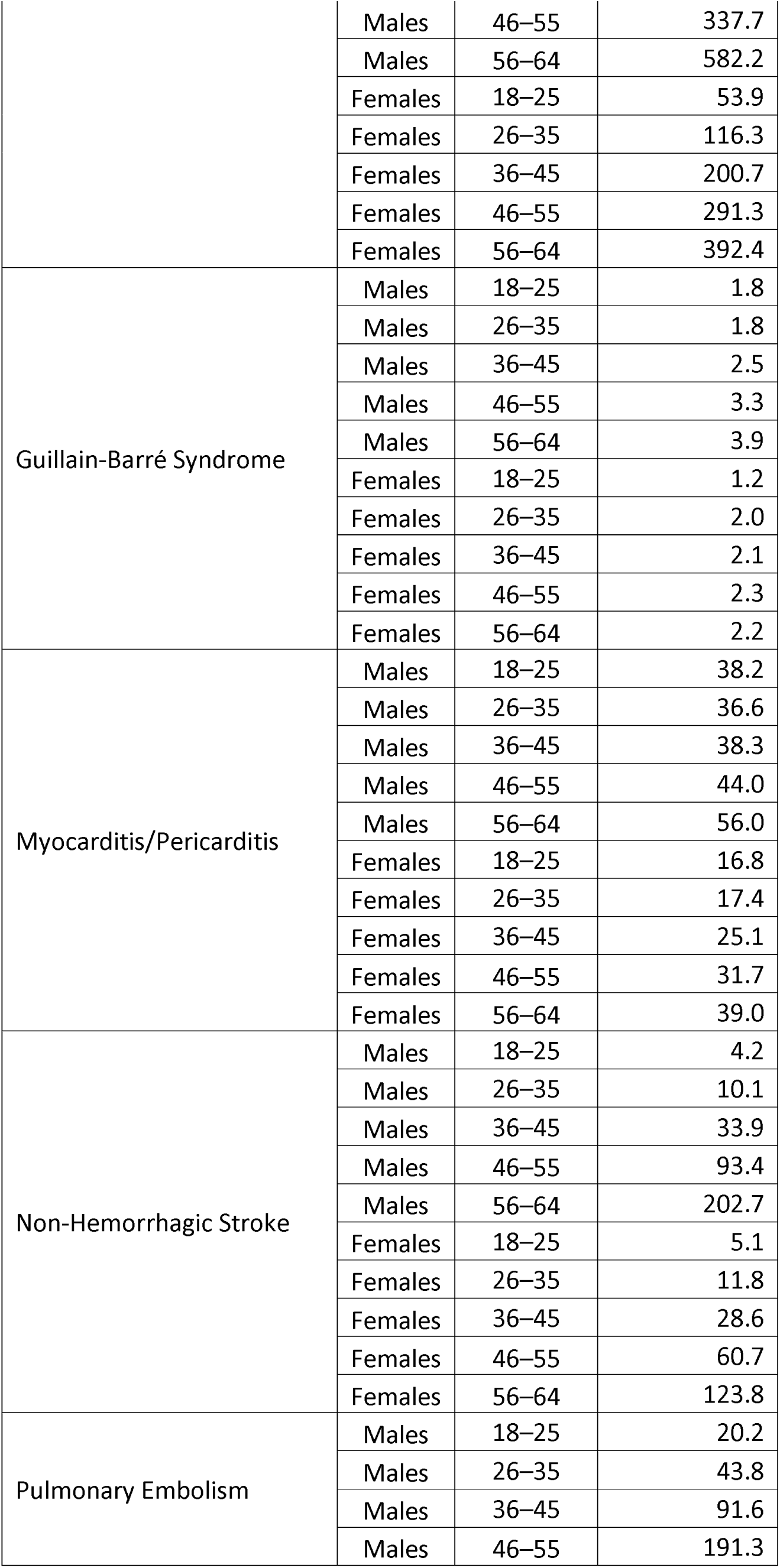
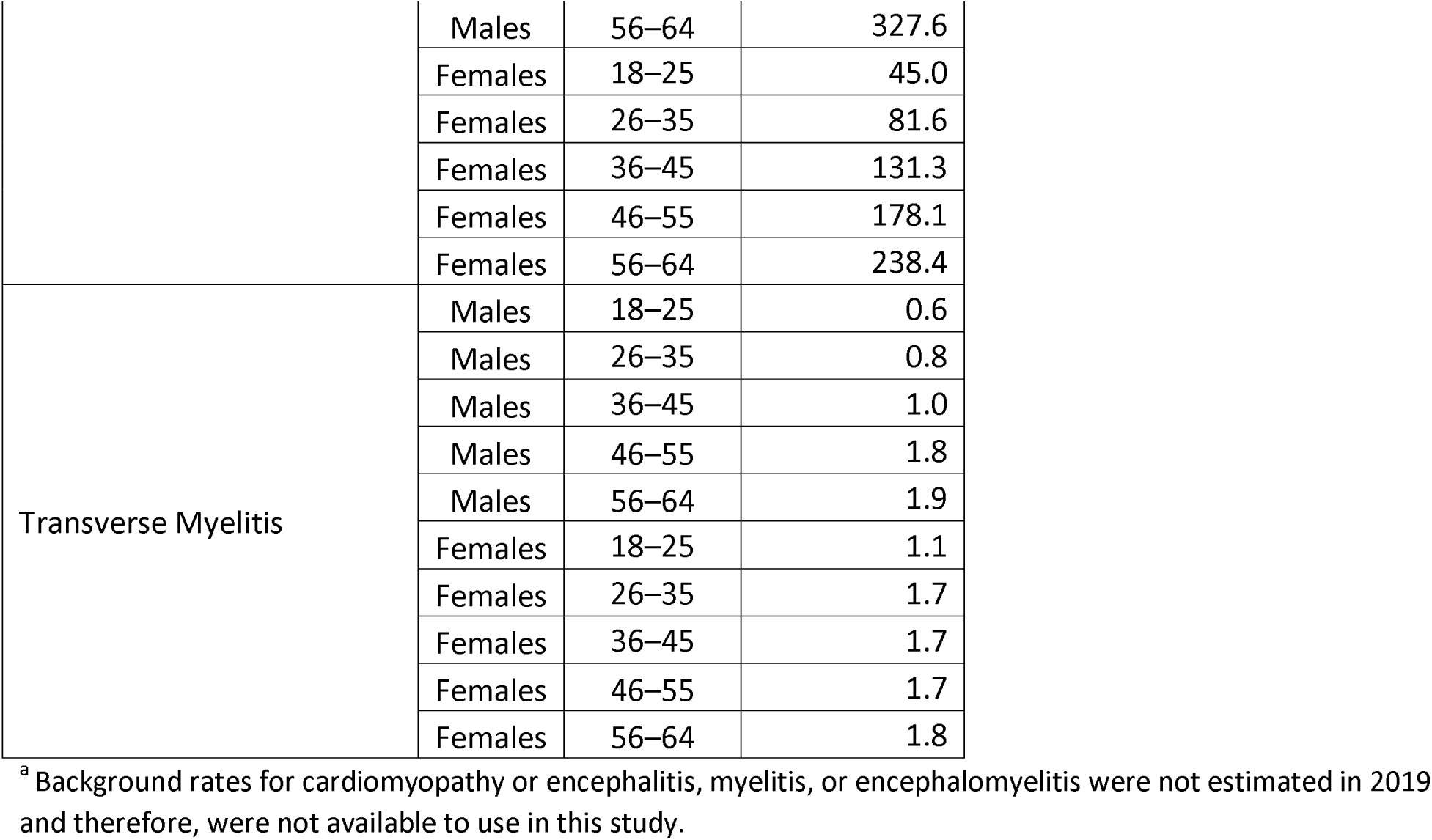
Sex- and age-specific 2019 background rates for each adverse event estimated from prior surveillance work using aggregated data from CVS Health, Carelon Research, and Optum commercial claims databases.

**Supplementary Table 3.**
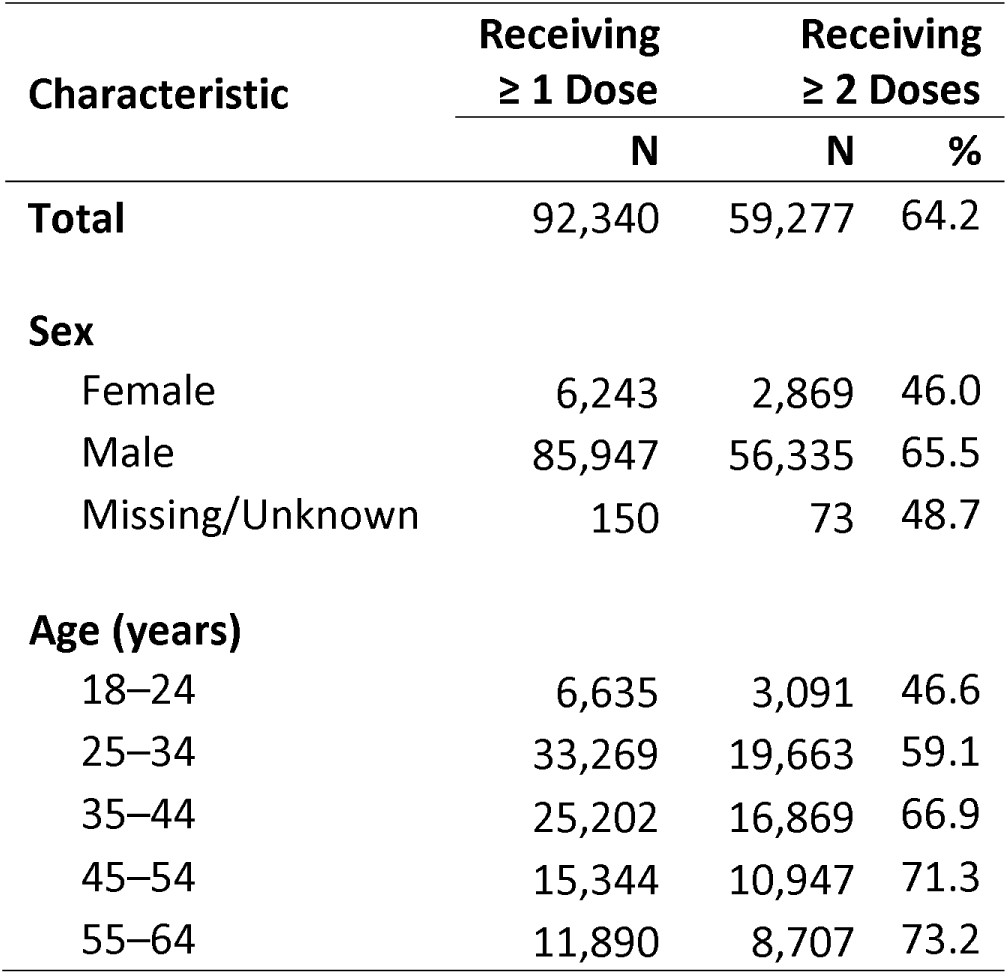
Number and percentage of people receiving a complete JYNNEOS vaccine series among U.S. adults aged 18–64 years who received at least one JYNNEOS dose.

## References

1. World Health Organization (WHO). Disease Outbreak News Multi-country monkeypox outbreak: situation update. 2022 [accessed October 1, 2024]; Available from: https://www.who.int/emergencies/disease-outbreak-news/item/2022-DON393.

2. Centers for Disease Control and Prevention (CDC). Archive Home: Ongoing 2022 Global Outbreak Cases and Data. [accessed October 1, 2024]; Available from: https://archive.cdc.gov/#/details?archive_url=https://archive.cdc.gov/www_cdc_gov/poxvirus/mpox/response/2022/index.html.

3. Centers for Disease Control and Prevention (CDC). Ongoing Clade II Mpox Global Outbreak. [accessed October 1, 2024]; Available from: https://www.cdc.gov/mpox/outbreaks/2022/index-1.html.

4. Tuttle, A., et al., Notes from the Field: Clade II Mpox Surveillance Update - United States, October 2023-April 2024. MMWR Morb Mortal Wkly Rep, 2024. 73(20): p. 474–476.

5. Centers for Disease Control and Prevention (CDC). CDC, Poxvirus, Monkeypox, Health Departments: Vaccine Considerations. 2022 [accessed October 1, 2024]; Available from: https://web.archive.org/web/20220811084742/https://www.cdc.gov/poxvirus/monkeypox/interim-considerations/overview.html.

6. Centers for Disease Control and Prevention (CDC). Interim Clinical Considerations for Use of Vaccine for Mpox Prevention in the United States. 2024 [accessed October 1, 2024]; Available from: https://www.cdc.gov/mpox/hcp/vaccine-considerations/vaccination-overview.html?CDC_AAref_Val=https://www.cdc.gov/poxvirus/mpox/interim-considerations/overview.html.

7. Food and Drug Administration (FDA). Full Prescribing Information - JYNNEOS. 2023 [accessed October 1, 2024]; Available from: https://www.fda.gov/media/131078/download.

8. Food and Drug Administration (FDA). Full Prescribing Information - ACAM200. 2024 [accessed November 8, 2024]; Available from: https://www.fda.gov/media/75792/download?attachment.

9. U.S. Food and Drug Administration (FDA). Key Facts About Vaccines to Prevent Monkeypox Disease. 2022 [accessed October 1, 2024]; Available from: https://www.fda.gov/vaccines-blood-biologics/vaccines/key-facts-about-vaccines-prevent-monkeypox-disease.

10. Liu, H., et al., Global perspectives on smallpox vaccine against monkeypox: a comprehensive meta-analysis and systematic review of effectiveness, protection, safety and cross-immunogenicity. Emerg Microbes Infect, 2024. 13(1): p. 2387442.

11. Duffy, J., et al., JYNNEOS Vaccine Safety Surveillance During the 2022 Mpox Outbreak Using the Vaccine Adverse Event Reporting System and V-safe, United States, 2022 to 2023. Sex Transm Dis, 2024. 51(8): p. 509–515.

12. Duffy, J., et al., Safety Monitoring of JYNNEOS Vaccine During the 2022 Mpox Outbreak - United States, May 22-October 21, 2022. MMWR Morb Mortal Wkly Rep, 2022. 71(49): p. 1555–1559.

13. Food and Drug Administration (FDA). Safety Surveillance of Vaccines Used for Mpox Prevention: Active Monitoring Master Protocol: Code List. 2022 [accessed October 1, 2024]; Available from: https://bestinitiative.org/wp-content/uploads/2023/01/Mpox-Vax-Safety-Monitoring-Protocol-Code-List-2022.xlsx.

14. Ulm, K., A simple method to calculate the confidence interval of a standardized mortality ratio (SMR). Am J Epidemiol, 1990. 131(2): p. 373–5.

15. Parker, J.D., et al., National Center for Health Statistics data presentation standards for rates and counts. Vital Health Stat, 2023. 2(200).

16. Food and Drug Administration (FDA). Background Rates of Adverse Events of Special Interest for COVID-19 Vaccine Safety Monitoring. 2022 [accessed October 8, 2024]; Available from: https://bestinitiative.org/wp-content/uploads/2022/08/Background-Rates-of-Adverse-Events-of-Special-Interest_for_COVID-19_Vaccine_Safety_Monitoring-All_Files.zip.

17. Office for Human Research Protections US Department of Health and Human Services. 45 CRF §46 2010 [accessed November 8, 2024]; Available from: https://www.hhs.gov/ohrp/regulations-and-policy/regulations/45-cfr-46/index.html.

18. Sharff, K.A., et al., Cardiac events following JYNNEOS vaccination for prevention of Mpox. Vaccine, 2023. 41(22): p. 3410–3412.

19. Delaney, K.P., et al., Strategies Adopted by Gay, Bisexual, and Other Men Who Have Sex with Men to Prevent Monkeypox virus Transmission - United States, August 2022. MMWR Morb Mortal Wkly Rep, 2022. 71(35): p. 1126–30.

